# Comparison of pandemic excess mortality in 2020-2021 across different empirical calculations

**DOI:** 10.1101/2022.05.09.22274863

**Authors:** Michael Levitt, Francesco Zonta, John P.A. Ioannidis

**Affiliations:** Department of Structural Biology, Stanford University, Stanford, CA 94305, USA; Shanghai Institute for Advanced Immunochemical Studies, ShanghaiTech University, Shanghai 201210, China; Departments of Medicine, of Epidemiology and Population Health, of Biomedical Data Science, and of Statistics, and Meta-Research Innovation Center at Stanford (METRICS), Stanford University, Stanford, CA 94305, USA

**Keywords:** COVID-19, mortality, excess mortality, modeling, epidemiology, excess deaths

## Abstract

Different modeling approaches can be used to calculate excess deaths for the COVID-19 pandemic period. We compared 6 calculations of excess deaths (4 previously published and two new ones that we performed with and without age-adjustment) for 2020-2021. With each approach, we calculated excess deaths metrics and the ratio R of excess deaths over recorded COVID-19 deaths. The main analysis focused on 33 high-income countries with weekly deaths in the Human Mortality Database (HMD at mortality.org) and reliable death registration. Secondary analyses compared calculations for other countries, whenever available. Across the 33 high-income countries, excess deaths were 2.0-2.8 million without age-adjustment, and 1.6-2.1 million with age-adjustment with large differences across countries. In our analyses after age-adjustment, 8 of 33 countries had no overall excess deaths; there was a death deficit in children; and 0.478 million (29.7%) of the excess deaths were in people <65 years old. In countries like France, Germany, Italy, and Spain excess death estimates differed 2 to 4-fold between highest and lowest figures. The R values’ range exceeded 0.3 in all 33 countries. In 16 of 33 countries, the range of R exceeded 1. In 25 of 33 countries some calculations suggest R>1 (excess deaths exceeding COVID-19 deaths) while others suggest R<1 (excess deaths smaller than COVID-19 deaths). Inferred data from 4 evaluations for 42 countries and from 3 evaluations for another 98 countries are very tenuous Estimates of excess deaths are analysis-dependent and age-adjustment is important to consider. Excess deaths may be lower than previously calculated.

**SIGNIFICANCE STATEMENT:** Excess deaths are a key metric for assessing the impact of a pandemic. They reflect the composite impact of deaths from infection, from indirect pandemic effects, and from the measures taken. Different modeling approaches can be used to calculate excess deaths for the COVID-19 pandemic. Here, we compare four previous calculations of excess deaths and two new ones that we performed with and without adjusting for changing age structure in the estimation. Proper age-adjustment results in substantial reduction in estimates of excess deaths for 2020-2021. While results from different calculation methods are correlated, the absolute differences in estimated excess deaths are very high in most countries. Extrapolations to countries without reliable death registration is extremely tenuous.

## INTRODUCTION

Many studies estimate excess deaths in specific locations, countries, regions, or worldwide during the COVID-19 pandemic.^1–4^ Excess deaths reflect a composite of deaths from SARS-CoV-2 infection plus indirect effects of the pandemic (e.g. health system strain) and measures taken.^5, 6^ It has been argued^7, 8^ that excess deaths are a more appropriate measure of impact than recorded COVID-19 deaths. Recorded COVID-19 deaths may be under- or over-counted in different time periods and locations^5^ and do not capture indirect effects of the pandemic and the measures taken. However, excess deaths calculations require modeling of the expected deaths that entails many assumptions and analytical choices. To obtain excess deaths estimates one needs to define a control (reference) pre-pandemic period, use some model for extrapolating expected deaths in the pandemic period and compare them against observed deaths. There are many different possibilities on how to select the pre-pandemic reference period and on how to model data and extrapolations.^9, 10^ A major analytical decision is whether to account for changes in the age structure of the population over time. With an aging population (particularly in high-income countries), mortality rates may increase over time, countering the anticipated decrease in mortality from better healthcare and overall human progress. These changes may be better addressed by considering age structure^9, 11–13^ rather than simply relying on regression trends of overall population data regardless of age.

Another pivotal dilemma in excess death calculations is what sources of data to use; and which countries are considered to have sufficiently reliable data. Death registration is sub-standard in most countries around the world: many deaths remain unrecorded.^14, 15^ Information on causes of death has flaws even in the most developed countries^16, 17^ while the pandemic generated new death coding challenges.^18^ Changes in deaths over time may be confounded by changes in death registration and recorded COVID-19 deaths depend on coding. Even population counts have uncertainty and this applies also to age-stratified estimates from different sources using different imputations to estimate age-stratified population for recent years. Even in high-income countries, most of them have not had a formal census performed for many years. Finally, several highly visible studies that attempted to calculate excess mortality world-wide,^1–4^ first estimated excess mortality in countries with trustworthy death data on all-cause mortality; then, they extrapolated across other countries worldwide, using the profile of various characteristics in these countries versus those with trustworthy data. Consequently, proper calculation of excess mortality in countries with most trustworthy mortality data has critical importance even for worldwide estimates.

Here, we compare the results of different evaluations that have attempted to calculate global excess mortality during the COVID-19 pandemic in 2020-2021.^1–4^ We focus primarily on high-income countries with the most reliable death registration systems and discuss the implications for extrapolations to a global level. We compared 4 widely publicized excess death calculations published in eLife, Economist, the Lancet, and by the World Health Organization (WHO)^1–4, 19, 20^ and included also our calculations. Our calculations explicitly explored what difference it would make to model deaths using separate death and population data for age strata. Besides raw excess mortality estimates, we focused on the ratio of excess mortality over recorded COVID-19 deaths. This ratio is critical in understanding whether excess mortality confers more information compared with just focusing on routinely recorded COVID-19 deaths. Ideally, one would like to see consistency in this ratio regardless of modeling and analytical choices, while large inconsistency would put the added value of excess mortality calculations in question.

## RESULTS

### Characteristics of the compared excess death calculations

Table 1 summarizes the main features of the compared excess death calculations. As shown, the evaluations differed substantially in defining the reference period, the choice of analytical model, exclusions, use of adjustments, eligibility criteria, data sources, extrapolations and imputations. The Economist methods used complex machine learning not described in very broad terms; the WHO and Lancet methods were convoluted and not all can be easily reproduced.

**Table 1.**
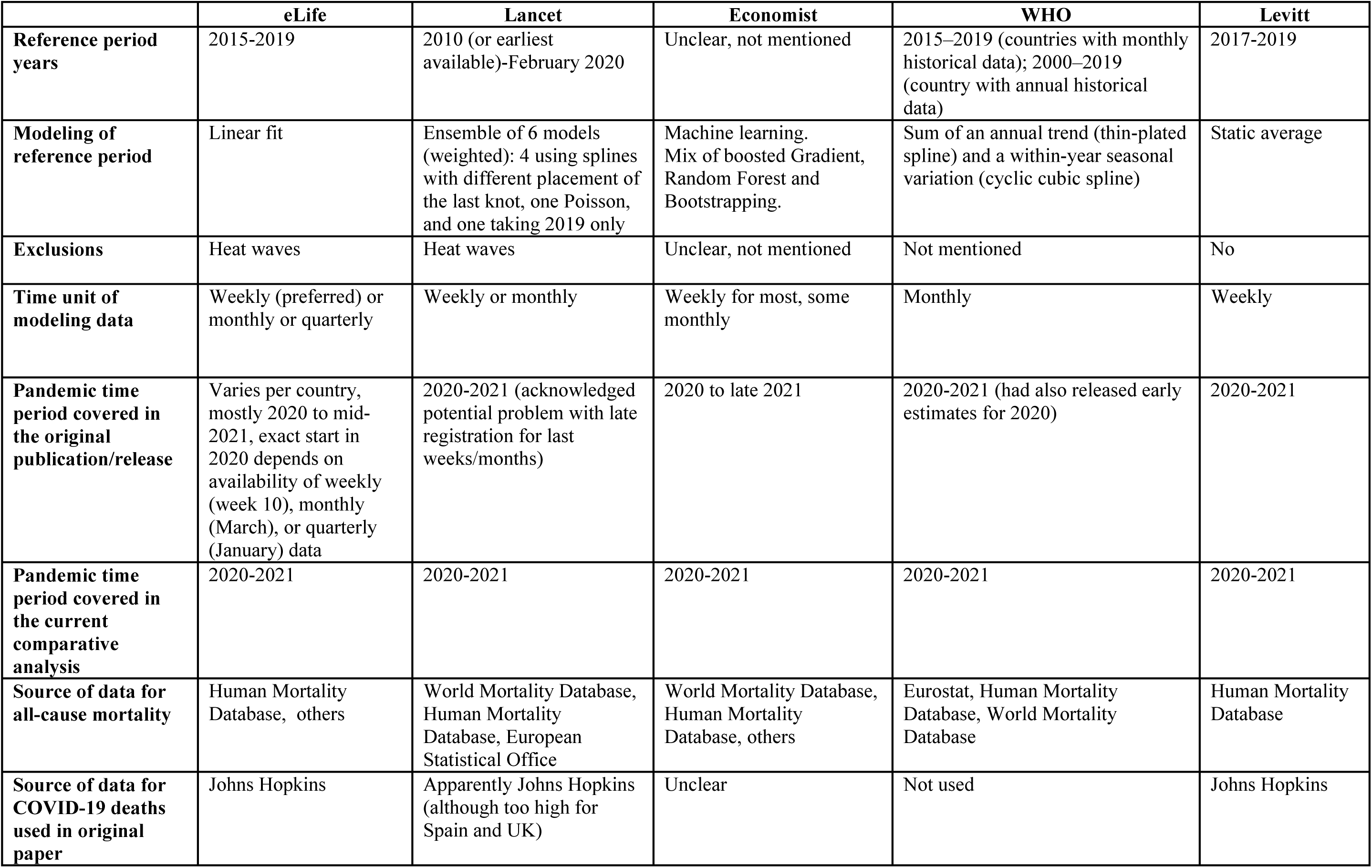
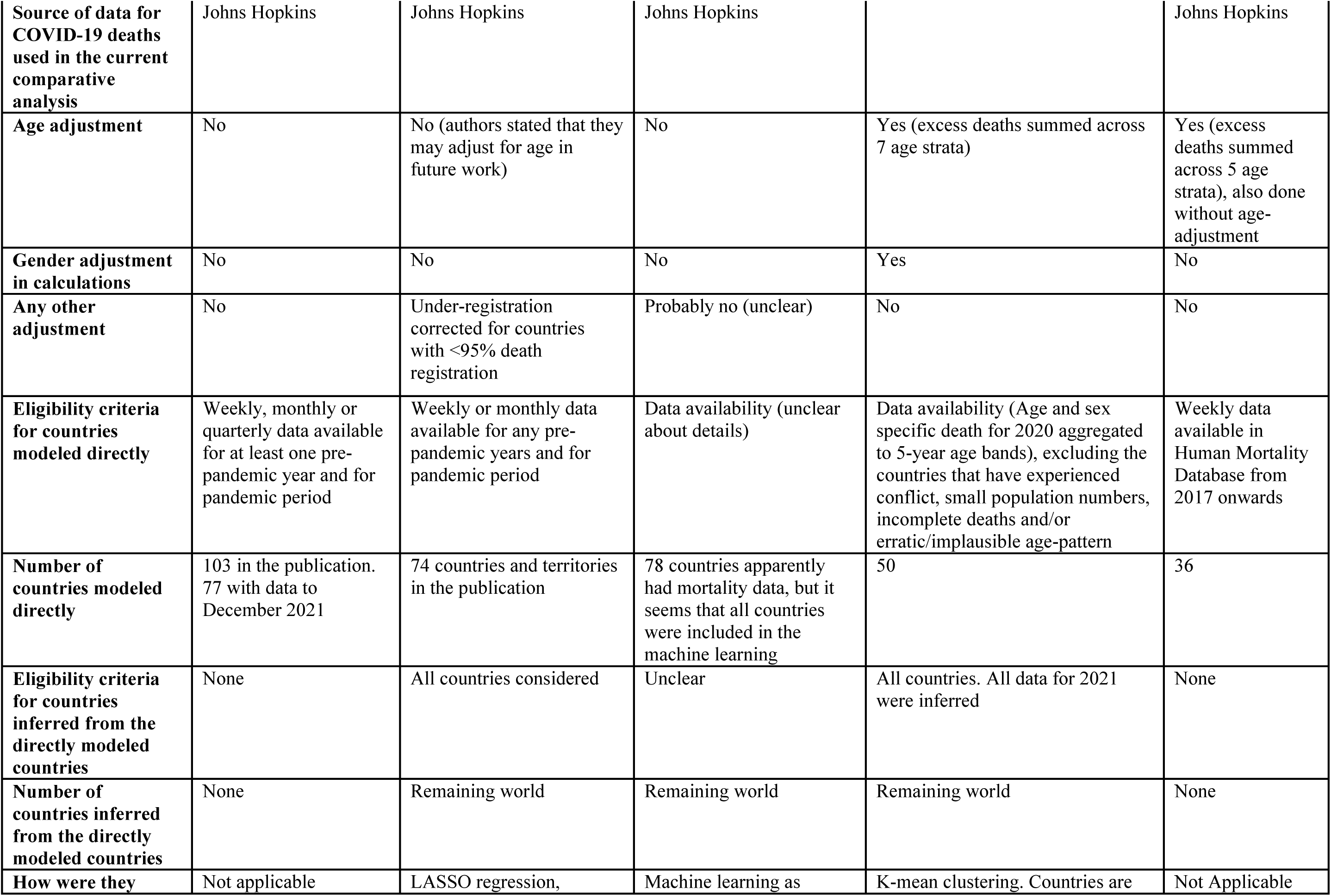
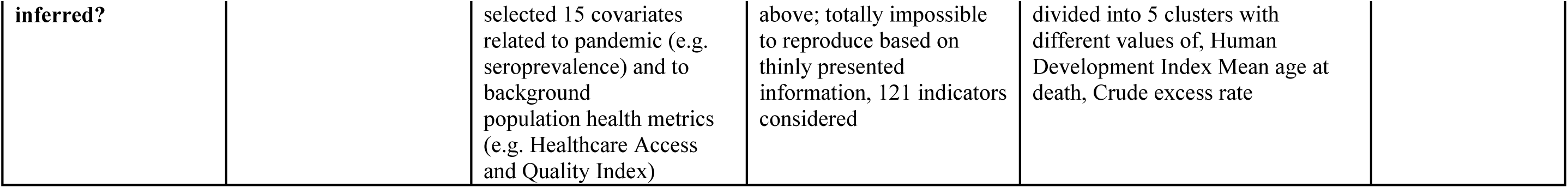
Main features of the construction of the compared evaluations of excess deaths.

### Comparison of excess deaths metrics in 33 countries with most-reliable data

33 countries were included in the main analysis, selected because they were high-income according to the World Bank, and they had detailed available weekly data on observed deaths in the three pre-pandemic years and also during 2020-2021 (Table 2). These countries had a total population of almost 1 billion and almost 20 million deaths in 2020-2021, of which 9.5% were recorded as COVID-19 deaths. The total of calculated excess deaths in these 33 countries ranged from 2.0 million (eLife) to 2.8 million (Lancet), with WHO (2.1 million) and Economist (2.2 million) being closer to eLife. Our own calculations without age adjustment gave a similar total (2.3 million), but with age-adjustment the excess deaths were only 1.6 million.

**Table 2.** Excess death estimates for 2020-2021 according to 6 evaluations in the 33 eligible countries*.

| Country | 2021<br>Population<br>(millions)<br>from HMD** | Two Years<br>Actual All-Cause Death<br>from HMD | Two Year<br>Excess Death<br>per eLife | Two Year<br>Excess<br>Death per<br>Economist | Two Year<br>Excess Death<br>per Lancet | Two Year<br>Excess<br>Death per<br>WHO | Two Year<br>Excess<br>Death per<br>Levitt Age-Adjusted | Two Year<br>Excess Death<br>per Levitt<br>Not Age-Adjusted |
| --- | --- | --- | --- | --- | --- | --- | --- | --- |
| Australia | 24.547 | 277,603 | -11,639 | -9,500 | -18,100 | -14,258 | -14,460 | -2,116 |
| Austria | 8.935 | 180,363 | 15,261 | 16,877 | 18,300 | 11,941 | 13,007 | 15,343 |
| Belgium | 11.494 | 239,201 | 20,613 | 23,364 | 32,800 | 17,919 | 13,958 | 19,036 |
| Canada | 36.108 | 586,135 | 13,474 | 23,548 | 43,700 | 22,018 | 21,829 | 37,938 |
| Chile | 17.960 | 263,154 | 38,894 | 38,094 | 37,200 | 38,698 | 31,640 | 45,021 |
| Croatia | 4.051 | 119,871 | 16,826 | 19,186 | 22,900 | 17,178 | 12,205 | 16,050 |
| Czechia | 10.730 | 269,137 | 41,480 | 43,942 | 49,100 | 37,040 | 34,079 | 43,262 |
| Denmark | 5.864 | 111,772 | 913 | 2,453 | 10,400 | 3,716 | -3,157 | 2,390 |
| Estonia | 1.332 | 34,559 | 3,172 | 3,774 | 5,630 | 3,374 | 2,675 | 3,346 |
| Finland | 5.548 | 112,800 | 2,662 | 4,469 | 8,780 | 2,858 | -716 | 4,345 |
| France | 65.467 | 1,297,407 | 78,910 | 97,390 | 155,000 | 81,849 | 57,767 | 96,831 |
| Germany | 82.533 | 2,005,701 | 88,446 | 113,242 | 203,000 | 194,987 | 54,740 | 128,557 |
| Greece | 10.711 | 274,725 | 24,177 | 25,269 | 25,400 | 19,394 | 20,515 | 29,551 |
| Hungary | 9.762 | 296,496 | 35,811 | 41,714 | 53,800 | 36,499 | 27,813 | 36,090 |
| Iceland | 0.362 | 4,640 | 50 | -35 | -314 | -10 | -142 | 11 |
| Israel | 9.293 | 99,437 | 7,203 | 7,967 | 9,280 | 6,178 | 3,201 | 5,421 |
| Italy | 59.630 | 1,454,193 | 167,816 | 190,872 | 259,000 | 160,800 | 115,690 | 166,373 |
| Latvia | 1.906 | 63,088 | 6,979 | 7,851 | 12,400 | 7,668 | 6,046 | 7,023 |
| Lithuania | 2.802 | 90,523 | 16,008 | 17,396 | 20,000 | 17,253 | 11,283 | 12,274 |
| Luxembourg | 0.635 | 9,106 | 57 | 314 | 1,070 | 69 | 109 | 171 |
| Netherlands | 17.479 | 339,242 | 28,495 | 33,017 | 45,500 | 29,213 | 17,969 | 32,020 |
| New Zealand | 5.013 | 67,586 | -2,787 | -2,566 | -872 | -2,678 | -4,118 | -1,826 |
| Norway | 5.408 | 82,491 | 1,101 | 1,986 | 742 | -100 | -2,994 | -182 |
| Poland | 38.482 | 998,284 | 157,247 | 171,806 | 214,000 | 157,531 | 149,722 | 182,454 |
| Portugal | 10.323 | 248,658 | 20,677 | 24,530 | 40,400 | 20,449 | 16,286 | 25,602 |
| Slovakia | 5.480 | 131,782 | 24,131 | 25,538 | 25,400 | 24,320 | 18,662 | 23,786 |
| Slovenia | 2.103 | 47,090 | 4,953 | 5,492 | 6,980 | 5,584 | 3,944 | 5,617 |
| South Korea | 51.631 | 621,862 | 7,529 | 6,967 | 4,630 | 6,289 | -30,286 | 33,417 |
| Spain | 47.511 | 948,016 | 102,991 | 115,685 | 162,000 | 103,935 | 68,720 | 95,964 |
| Sweden | 10.408 | 184,326 | 9,926 | 11,976 | 18,100 | 11,253 | -367 | 3,666 |
| Switzerland | 8.688 | 146,969 | 11,394 | 13,539 | 15,500 | 8,247 | 5,640 | 10,139 |
| United Kingdom | 67.145 | 1,353,941 | 136,795 | 148,889 | 169,000 | 148,896 | 87,307 | 125,716 |
| United States | 329.995 | 6,849,500 | 961,032 | 1,017,655 | 1,130,000 | 932,460 | 871,295 | 1,116,088 |
| TOTALS | 969.336 | 19,809,658 | 2,030,597 | 2,242,701 | 2,780,726 | 2,110,570 | 1,609,862 | 2,319,376 |

HMD , Human Mortality Database short-term mortality fluctuation file stmf.csv downloaded from https://www.mortality.org/Public/STMF/Outputs/stmf.csv on 1-May-2022.
Data is given for full two-year period 1-Jan-2020 to 31-Dec-2021 for all countries except for Australia to 2021 week 48 and Canada to 2021 week 48. Because we use a standard year of 365.25 days (52.1786 weeks), the two years 2020 & 2021 are 104.357 weeks. The HMD Total Deaths, Expected Deaths and Excess Deaths for Australia and Canada are all smaller than they would be if data for these two locations were no delayed.
Data in the HMD are summed over weeks available. This means that both the Total Deaths and Population are incomplete for Australia (to week 47) and Canada (to week 48).

There were very large differences across countries and this was evident also when excess deaths were estimated as a proportion above the observed deaths, E_p_ (Supplementary Table 1), deaths per million population, E_m_ (Supplementary Table 2) and ratio of excess deaths over recorded COVID-19 deaths, R (Table 3) metrics. Australia, New Zealand, and in some analyses also Iceland showed no excess deaths at all, even without age-adjustment. With age-adjustment, however, in our calculations several other countries such as Norway, Finland, Denmark, Sweden, and South Korea also showed no excess deaths. In the WHO age-adjusted calculations, no excess deaths were seen in Australia, New Zealand, Iceland, and Norway. Among countries with excess deaths in all analyses, the excess death estimate differed by 2-4 times between the highest (non-age-adjusted) and lowest (age-adjusted) estimates in countries like France, Germany, Italy, and Spain. Age-adjustment led to modest reductions of estimates for the USA.

**Table 3.** Ratio of excess deaths over recorded COVID-19 deaths for 33 countries.

| Country | Reported COVID-19 Deaths from OWID | Excess Death/Reported per eLife | Excess Death/Reported Economist | Excess Death/Reported per Lancet | Excess Death/Reported per WHO | Excess Death/Reported Age-Adjusted | Excess Death/Reported per Levitt Not Age-Adjusted |
| --- | --- | --- | --- | --- | --- | --- | --- |
| Australia | 2,253 | -5.17 | -4.22 | -8.03 | -6.33 | -6.42 | -0.94 |
| Austria | 13,733 | 1.11 | 1.23 | 1.33 | 0.87 | 0.95 | 1.12 |
| Belgium | 28,331 | 0.73 | 0.82 | 1.16 | 0.63 | 0.49 | 0.67 |
| Canada | 30,570 | 0.44 | 0.77 | 1.43 | 0.72 | 0.71 | 1.24 |
| Chile | 39,115 | 0.99 | 0.97 | 0.95 | 0.99 | 0.81 | 1.15 |
| Croatia | 12,538 | 1.34 | 1.53 | 1.83 | 1.37 | 0.97 | 1.28 |
| Czechia | 36,129 | 1.15 | 1.22 | 1.36 | 1.03 | 0.94 | 1.20 |
| Denmark | 3,267 | 0.28 | 0.75 | 3.18 | 1.14 | -0.97 | 0.73 |
| Estonia | 1,932 | 1.64 | 1.95 | 2.91 | 1.75 | 1.38 | 1.73 |
| Finland | 1,714 | 1.55 | 2.61 | 5.12 | 1.67 | -0.42 | 2.53 |
| France | 123,805 | 0.64 | 0.79 | 1.25 | 0.66 | 0.47 | 0.78 |
| Germany | 111,925 | 0.79 | 1.01 | 1.81 | 1.74 | 0.49 | 1.15 |
| Greece | 20,790 | 1.16 | 1.22 | 1.22 | 0.93 | 0.99 | 1.42 |
| Hungary | 39,186 | 0.91 | 1.06 | 1.37 | 0.93 | 0.71 | 0.92 |
| Iceland | 37 | 1.35 | -0.95 | -8.49 | -0.27 | -3.82 | 0.29 |
| Israel | 8,243 | 0.87 | 0.97 | 1.13 | 0.75 | 0.39 | 0.66 |
| Italy | 137,402 | 1.22 | 1.39 | 1.88 | 1.17 | 0.84 | 1.21 |
| Latvia | 4,570 | 1.53 | 1.72 | 2.71 | 1.68 | 1.32 | 1.54 |
| Lithuania | 7,387 | 2.17 | 2.35 | 2.71 | 2.34 | 1.53 | 1.66 |
| Luxembourg | 915 | 0.06 | 0.34 | 1.17 | 0.08 | 0.12 | 0.19 |
| Netherlands | 20,999 | 1.36 | 1.57 | 2.17 | 1.39 | 0.86 | 1.52 |
| New Zealand | 51 | -54.65 | -50.31 | -17.10 | -52.51 | -80.75 | -35.81 |
| Norway | 1,305 | 0.84 | 1.52 | 0.57 | -0.08 | -2.29 | -0.14 |
| Poland | 97,054 | 1.62 | 1.77 | 2.20 | 1.62 | 1.54 | 1.88 |
| Portugal | 18,955 | 1.09 | 1.29 | 2.13 | 1.08 | 0.86 | 1.35 |
| Slovakia | 16,635 | 1.45 | 1.54 | 1.53 | 1.46 | 1.12 | 1.43 |
| Slovenia | 5,589 | 0.89 | 0.98 | 1.25 | 1.00 | 0.71 | 1.01 |
| South Korea | 5,625 | 1.34 | 1.24 | 0.82 | 1.12 | -5.38 | 5.94 |
| Spain | 89,405 | 1.15 | 1.29 | 1.81 | 1.16 | 0.77 | 1.07 |
| Sweden | 15,310 | 0.65 | 0.78 | 1.18 | 0.74 | -0.02 | 0.24 |
| Switzerland | 12,217 | 0.93 | 1.11 | 1.27 | 0.68 | 0.46 | 0.83 |
| United Kingdom | 148,737 | 0.92 | 1.00 | 1.14 | 1.00 | 0.59 | 0.85 |
| United States | 827,887 | 1.16 | 1.23 | 1.36 | 1.13 | 1.05 | 1.35 |
| TOTAL OR<br>MEDIAN | 1,883,611 | 1.09 | 1.22 | 1.36 | 1.00 | 0.71 | 1.15 |

OWID refers to Our World in Data master COVID-19 file down loaded from https://covid.ourworldindata.org/data/owid-covid-data.csv on 22-Apr-2022.

The pairwise correlations between eLife, Economist, WHO and our two calculations (with and without age-adjustments) were consistently extremely high for R (all r>=0.977). The same picture was seen largely for these 5 calculations for E_p_ (all r>=0.930) and for E_m_ (all r>=0.951). The Lancet calculations had modestly lower correlation with the other 4 evaluations (range r=0.808-0.841 for R, range r=0.835-0.919 for E_p_, 0.917-0.958 for E_m_). The correlations for R between different evaluations, however, were modest/poor when Australia and New Zealand (that were outliers with very negative values of R) were excluded, (range -0.386 to 0.657). The two other metrics E_p_ (range, 0.794-0.989) and E_m_ (range 0.904-0.996) show good correlation values even when these outliers are excluded.

Even when correlations were high, given the substantial differences in the absolute estimates of excess deaths with the different calculations in each country, the range of R values was always large (Table 3). The R values’ range always exceeded 0.3 in all 33 countries. In 16 of 33 countries, the range of R across different evaluations exceeded 1, i.e. it was as large as the number of recorded COVID-19 deaths itself. 6 countries had consistently R >1, i.e. more excess deaths than recorded COVID-19 deaths, with all empirical evaluations. Conversely, only 2 countries, Australia and New Zealand had consistently R<1. The large majority of countries (25 of 33) had some calculations suggesting R>1 (i.e. excess deaths greater than COVID-19 deaths) and others suggesting R<1 (excess deaths smaller than COVID-19 deaths).

Table 4 shows the break-down of excess deaths per age stratum in each country for our age-adjusted analyses. In total, 0.478 of the 1.609 million excess deaths were in people <65 years old (29.7%), but the percentage varied widely across countries. 30 of 33 countries had death deficit for children 0-14 years old (all, except for Iceland, Luxembourg, and Netherland that also had minimal excess deaths in children) and overall across all 33 countries there was a death deficit of 7,737 deaths for children 0-14 years old. 10 countries had death deficit even for people <65 years old. Conversely, for 4 countries, more than 25% of the excess deaths was in people <65 years old (Canada 45.5%, USA 41.6%, Chile 37.8%, UK 28.7%).

**Table 4.** Excess deaths per age strata in the 33 countries of the main analysis.

| Country | Mean HMD<br>2020 & 2021<br>Population<br>(millions) | Excess<br>deaths<br>in 0-14<br>years | Excess<br>deaths<br>in 15-64<br>years | Excess<br>deaths<br>in 65-74<br>years | Excess<br>deaths<br>in 75-84<br>years | Excess<br>deaths<br>in >85<br>years | Excess<br>death for<br>all ages | Percentage<br>of excess<br>deaths <65<br>years old |
| --- | --- | --- | --- | --- | --- | --- | --- | --- |
| Australia | 24.547 | -151 | -1,196 | -1,802 | -5,485 | -5,825 | -14,460 | No excess |
| Austria | 8.935 | -55 | 1,812 | 1,449 | 5,673 | 4,129 | 13,007 | 13.5% |
| Belgium | 11.494 | -307 | 675 | 2,707 | 4,071 | 6,812 | 13,958 | 2.6% |
| Canada | 36.108 | 1,113 | 8,894 | 4,429 | 4,558 | 2,835 | 21,829 | 45.8% |
| Chile | 17.960 | -857 | 12,810 | 7,551 | 7,592 | 4,545 | 31,640 | 37.8% |
| Croatia | 4.051 | -25 | 1,192 | 3,639 | 4,196 | 3,203 | 12,206 | 9.6% |
| Czechia | 10.730 | -144 | 4,261 | 9,906 | 11,614 | 8,442 | 34,079 | 12.1% |
| Denmark | 5.864 | -21 | -882 | -194 | -1,643 | -417 | -3,157 | No excess |
| Estonia | 1.332 | -9 | 530 | 538 | 741 | 875 | 2,675 | 19.5% |
| Finland | 5.548 | -35 | -350 | 308 | -652 | 12 | -716 | No excess |
| France | 65.467 | -562 | -3,076 | 13,541 | 8,666 | 39,198 | 57,767 | No excess |
| Germany | 82.533 | -143 | 12,197 | 10,066 | 34,161 | -1,541 | 54,740 | 22.0% |
| Greece | 10.711 | -70 | 3,569 | 4,710 | 3,500 | 8,805 | 20,515 | 17.1% |
| Hungary | 9.762 | -94 | 4,593 | 10,362 | 8,676 | 4,277 | 27,813 | 16.2% |
| Iceland | 0.362 | 19 | 6 | -14 | -139 | -14 | -142 | ** |
| Israel | 9.293 | -269 | 203 | 1,168 | 668 | 1,431 | 3,201 | -2.1% |
| Italy | 59.630 | -568 | 12,066 | 21,888 | 40,081 | 42,223 | 115,690 | 9.9% |
| Latvia | 1.906 | -49 | 1,082 | 1,248 | 2,254 | 1,511 | 6,046 | 17.1% |
| Lithuania | 2.802 | -56 | 2,584 | 2,403 | 3,567 | 2,785 | 11,283 | 22.4% |
| Luxembourg | 0.635 | 40 | -58 | -64 | 1 | 189 | 109 | No excess |
| Netherlands | 17.479 | 14 | 1,241 | 3,566 | 7,175 | 5,973 | 17,969 | 7.0% |
| New Zealand | 5.013 | -62 | -512 | -797 | -1,301 | -1,446 | -4,118 | No excess |
| Norway | 5.408 | -63 | -452 | -433 | -1,540 | -506 | -2,994 | No excess |
| Poland | 38.482 | -574 | 19,293 | 47,295 | 39,417 | 44,291 | 149,722 | 12.5% |
| Portugal | 10.323 | -110 | 1,959 | 2,877 | 4,277 | 7,283 | 16,286 | 11.4% |
| Slovakia | 5.480 | -36 | 3,467 | 6,293 | 6,211 | 2,728 | 18,663 | 18.4% |
| Slovenia | 2.103 | -13 | -49 | 691 | 1,418 | 1,897 | 3,944 | No excess |
| South_Korea | 51.631 | -774 | -431 | -7,474 | -10,335 | -11,272 | -30,286 | No excess |
| Spain | 47.511 | -203 | 9,788 | 13,194 | 14,342 | 31,598 | 68,720 | 13.9% |
| Sweden | 10.408 | -41 | -682 | 2 | -422 | 776 | -367 | No excess |
| Switzerland | 8.688 | -16 | 175 | 492 | 1,075 | 3,914 | 5,640 | 2.8% |
| United_Kingdom | 67.145 | -762 | 25,852 | 19,276 | 20,476 | 22,465 | 87,307 | 28.7% |
| United_States | 329.995 | -2,858 | 365,676 | 216,688 | 169,623 | 122,167 | 871,295 | 41.6% |
| TOTAL | 969.336 | -7,737 | 486,236 | 395,508 | 382,515 | 353,341 | 1,609,862 | 29.7% |

\*\*No excess overall, small excess in >65 years old.

### Other countries

eLife, Economist, Lancet, and WHO had estimates of excess deaths in 42 additional countries (Supplementary Table 3). These countries had a total population of almost 1.4 billion, the total reported COVID-19 deaths were almost 2.3 million and the excess death estimates were about double with all 4 evaluations (4.7 million per eLife, 4.8 million per Economist, 5.5 million per Lancet, 4.4 million per WHO) with higher correlations between eLife, Economist and WHO and more modest correlations with Lancet for all three excess death metrics. For several countries, differences across evaluations were very large. Two countries had death deficits by some evaluations, but not with others (Singapore range -1,770 to 1,776 and Japan range -19,469 to 111,000).

Another 98 countries (total population 5.3 billion) had excess death values obtained only per Economist, Lancet and WHO (Supplementary Table 4) for a total of 10.7, 9.8, and 7.9 million, respectively (6-9 times higher than recorded COVID-19 deaths). The correlation of the calculations was modest for all three metrics (r=0.499-0.570, r=0.552-0.662, r=0.530-0.801 for E_p_, E_m_, and R, respectively). Very large differences across different calculations for the same country were very common. For 11 countries, there was an estimated death deficit based on some calculation but not with all 3. For China, the range was extreme (452,669 excess deaths per Economist; -52,064 deficit per WHO).

## DISCUSSION

Across 33 high-income countries with total population of approximately 1 billion and highly thorough death registration, different empirical estimates of excess deaths in 2020-2021 ranged widely from 1.6 million, i.e. substantially fewer than the 1.9 million recorded COVID-19 deaths, to 2.8 million, almost a million more. Countries with highest estimates of excess deaths were more or less the same in all evaluations; and countries with the most favorable picture performed well across the different evaluations. However, large differences emerged in the magnitude of the estimates for each country. The largest divergence was produced by whether age adjustment was used in calculating excess deaths. Age-adjusted estimates were lower. They are more appropriate, since COVID-19 has impressive age-related risk gradient for death.^20, 21^ Modest changes in age structure, in particular with aging populations, may produce major differences in estimates. With age-adjustment, several high-income countries showed no or minimal excess deaths; and for many others, the estimates of excess deaths markedly decreased. However, even age-adjusted excess death estimates differed substantially depending on the choice of data source for age-stratified population in the recent years as well as modeling choices.

Our age-adjusted analyses also revealed large differences across countries in the proportion of excess deaths accounted by non-elderly age strata. A few countries had more than a quarter of the excess deaths in people <65 years old. There are several possible factors that may explain this pattern. Adverse risk profile of the non-elderly populations, including high prevalence of obesity; inequalities and disadvantaged populations without good health care; and increased fatalities due to opioid overdose and other non-COVID-9 causes of excess death may explain in part the USA pattern. Also for Canada, USA and UK, many deaths in elderly people may have occurred in long-term care residents with very limited life expectancy. Deaths from SARS-CoV-2 among patients with limited life expectancy result in no excess deaths if the time window assessed after their infection is shorter than their life expectancy.^23^

The ratio R of excess deaths over recorded COVID-19 deaths varied substantially across different evaluations. Only 5 countries in Eastern Europe and USA had consistently R >1, i.e. more excess deaths than recorded COVID-19 deaths. Conversely, only Australia and New Zealand had consistently R<1. R values varied widely for all countries. In most countries, the uncertainty in the range of excess death estimates exceeded the number of recorded COVID-19 deaths itself. This large variability questions to what extent excess deaths can give much better insights on the total pandemic toll than COVID-19 recorded deaths.^7^ Excess death calculations are dependent on how they are calculated. In some cases, like Eastern Europe, they can tell that COVID-19 deaths have been undercounted and/or the numbers of other deaths have escalated during the pandemic. However, they cannot differentiate the relative contribution of these two factors nor can they give a precise estimate of either or their combination. In a few countries with limited SARS-CoV-2 deaths they can be reassuring that indirect pandemic effects and measures did not escalate fatalities, at least during 2020-2021. However, for most countries, excess death calculations are so model-dependent that they should be seen with great caution.

Besides the high-income countries with meticulous weekly death registration data, excess death calculations for the rest of the world are very tenuous exercises. For 42 countries where eLife, Economist, Lancet and WHO generated estimates, on average excess deaths were ∼2-times the COVID-19 recorded deaths and among another 98 countries where Economist and Lancet provided estimates, overall R was 6-9. However, given the low reliability of the data, the immense uncertainty surrounding these estimates cannot be overstated. More importantly, these calculations offer no causal insights. Excess deaths may be due to the virus, the indirect pandemic effects, or/and disruptive measures taken, even more so in countries with very frail healthcare systems, widespread poverty, and/or even high rates of hunger. The 2020-2021 crisis may have indeed resulted in many deaths, but causes may be very complex.

For some of these additional countries, calculations were run based on available mortality data. Even then, in most cases death registration is unreliable and the impact of changes in death registration during the pandemic compound any calculation. Perhaps age-adjustment would also lead to different estimates in these countries, as for the 33 most data-reliable countries. Even for these 33 countries, population estimates (including age-stratified population counts) are projected with different methods and typically no formal population census has been performed for several years. This adds further uncertainty to excess death calculations that are highly susceptible to minor differences especially in the population of elderly strata. For most countries, excess death calculations are indirectly imputed from the countries with mortality data. The methods employed by the Economist are not described in sufficient detail to allow probing validity and reproducibility. Lancet and WHO calculations provide more elaboration, mostly proving their complexity. The uncertainty in excess death estimates apparently far exceeds the width of published confidence intervals.

Hence, extrapolations from the 33 main analysis countries to other countries need to be extremely cautious. Among the total excess deaths, deaths due specifically to viral infection in the other countries may be proportionally far less. Other countries have far smaller percentage of elderly people and very few nursing home residents. Conversely, they have far more frail health systems, societies, and economies. Therefore, probably indirect effects of the pandemic and measures were perhaps more important contributors to total excess deaths than SARS-CoV-2 infection.

Nepomuceno et al. have also assessed the impact of different analytical choices on excess death calculations for 2020.^8^ They find modest differences among different approaches in countries with reliable death registration. Age-stratification has been also to be important in previous assessments for specific countries.^24, 25^ Germany is a classic example. Our age-adjusted estimate is 55,000 excess deaths, while without age-adjustment we calculated 129,000 excess deaths and Lancet calculated 203,000 excess deaths – compared with 111,000 COVID-19 reported deaths. Baum^23^ calculated only 22,000 excess deaths after age adjustment, while Koenig^24^ without age adjustment found more excess deaths than recorded COVID-19 deaths. In Germany, the number of people aged >80 years increased from 4.6 million in 2016 to 5.8 million in 2020, so consideration of age is crucial.^11^ Further diversity stems from whether modeling anticipates increasing life expectancy (decreasing mortality rate) over time.^11^ However, this approach may calculate spuriously high excess deaths: calculations assume a desired mortality rate lower than even attained in reality in the past. Medical and overall progress cannot guarantee continuing to decrease overall mortality in high-income countries, especially in old, frail people. In fact, care of such people may have deteriorated over time (e.g. with privatization and deterioration of long-term care) and the pandemic brought this to light.^26^ Moreover, data from much longer-term periods of observation suggest multiple, overlapping, complex long-term trends in winter mortality.^27^ Finally, some other models may diverge in their calculations, if they exclude certain periods. E.g., the popular Euromomo model using a Serfling model in its core but excludes weeks of high influenza activity from the modeling: thus it generates high excess death estimates.^28^

Some other caveats should be discussed. First, finer adjustment for age (e.g. in more narrow age bins), and more comprehensive adjustment for other factors (e.g. gender, frailty, long-term care facility residence, comorbidities) may be able to offer even more accurate estimates of excess deaths. Second, even in countries with reliable data, some deaths are registered with delays, although with >4 months after the end of 2021 our main analyses for the 33 countries should not be affected much. Third, there can be debate on whether/how natural disasters and wars should be excluded. Fourth, long-term effects of both the pandemic and measures taken on healthcare, other aspects of health, education, society, and economy remain uncaptured in the 2020-2021 window. Comparisons of different approaches to excess death calculations should continue for longer periods of follow-up, also in the post-pandemic endemic phase. The boundaries between pandemic and endemic phase can be debated.^29^ Regardless, the relative performance of different countries and their excess death ranking may change substantially over time. Sixth, analyzing specific causes of death may be informative, but suffers from major misclassification even in high-income countries.

Acknowledging these caveats, our analyses map the magnitude and uncertainty of excess deaths during 2020-2021. In countries with reliable data, age-adjustment suggests that excess deaths is lower than what has been previously published in calculations without age-adjustment. Excess death calculations convey some broad picture, especially for countries that fared very well or very poorly. Large differences in the impact of a pandemic across countries has been seen also in previous pandemics^30, 31^ for reasons that often remain largely unexplained. In depth assessments with death certificate audits, medical record audits, and autopsies may yield more granular insights about deaths and their causes, but these approaches also have limitations and feasibility challenges. For most countries worldwide, the tremendous uncertainty in the sparse data and indirect inferences should be mostly a call for improving the completeness and accuracy of death registration and investment in more rigorous demography infrastructures in the future.

## MATERIALS AND METHODS

### Compared published excess mortality estimates

We considered four previously published pandemic excess mortality evaluations^1–4, 18, 19^ that have calculated both country-specific and global estimates and which use diverse methods to extrapolate from pre-pandemic reference periods to the pandemic period.

Karlinsky and Kobak published their evaluation in eLife in mid-2021;^1^ the Economist team released their estimates in late 2021;^2^ the COVID-19 Excess Mortality Collaborators published their estimates in the Lancet in early 2022 considering the two-year period 2020-2021;^3^ and WHO released in May 2022 its updated estimates covering the same two year period 2020-2021. We call these four evaluations for convenience eLife, Economist, Lancet, and WHO respectively. Both the eLife and Economist models allow updating of excess mortality estimates over time and we have used the Our World in Data resource^18, 19^ that includes such updates. We used the estimates of excess mortality for the 2020-2021 two-year period for all 4 evaluations to maximize comparability. The three evaluations used also different sources for capturing the recorded numbers of COVID-19 deaths, which resulted in mostly minor discrepancies. Again, to maximize comparability, we used the same set of recorded numbers of COVID-19 for comparing against the excess mortality estimates of each evaluation: this set is identical to the set used by the Lancet evaluation, with the exception of Spain and UK where the Lancet numbers of recorded deaths were too high by 10% and 16%, respectively (perhaps due to clerical error) and where we used the Johns Hopkins data instead.^32^

The reported cumulative counts of COVID-19 deaths are taken from the Johns Hopkins Repository as reported by Our World in Data. The same values are reported by two of the four methods we use: eLife and Economist. Lancet reported values in https://ghdx.healthdata.org/sites/default/files/record-attached-files/IHME_EM_COVID_19_2020_2021_DATA_Y2022M03D10.CSV are sometimes quite different. Specifically for Spain, United Kingdom, Kazakhstan, Mexico, Russia, Georgia & Tajikistan where discrepancies are over 10% and some times as large as 115% (Russia is 651,000 in Lancet and 302,671 in OWID).

For each of the 4 evaluations, we extracted information on the following methodological features: reference period selected; modeling of reference period (static average, linear, spline, Poisson seasonality, other); exclusion of heat waves, wars, natural disasters, other; unit of modeling data (week, month, quarter, other); pandemic time period covered in the original publication/release; source of data for all-cause mortality; source of data for COVID-19 deaths used in original analysis; age and/or gender adjustment in calculations of excess deaths (if yes, how); any other adjustment in the calculations (if yes, specify); eligibility criteria and number of countries modeled directly; eligibility criteria and number of countries inferred from the directly modeled countries; and how these were inferred. Details on the data sources and modeling methods for these 4 evaluations can be found in references 1-4, 18 and 19.

### Evaluation considering age strata in the calculations

We performed also our own calculation of excess deaths focused on considering the impact of age-adjustment on the calculations. We considered three pre-pandemic years (2017 to 2019) as the reference period. We used the following age strata: 0-14, 15-64, 65-75, 75-85, and >85 years old. For each age stratum, we obtained the average mortality, the number of deaths per million for the population of the specific age stratum. Then we extrapolated to the two pandemic years, again correcting for the population size in the specific age stratum. Finally, expected deaths were summed across the population strata. An illustrative worked example of the age-adjusted calculations is shown in Figure 1 using the data for Germany. This analytical approach corrects for changes in total population of a country over time, as well as changes in the proportion of elderly people. If the total population and/or the proportion of older people is increasing in recent years over time, the expected deaths by the age-adjusted scheme will be more compared to analyses that do not consider population changes and age-stratification. The inverse will happen, if the total population and/or the proportion of older people is decreasing in recent years over time. We used the Human Mortality Database (https://www.mortality.org),33 specifically the Short Term Mortality Fluctuations file (https://www.mortality.org/Public/STMF/Outputs/stmf.csv) that includes weekly all-cause deaths and mortality values allowing the population size to be calculated as their ratio for each week.

**Figure 1:**
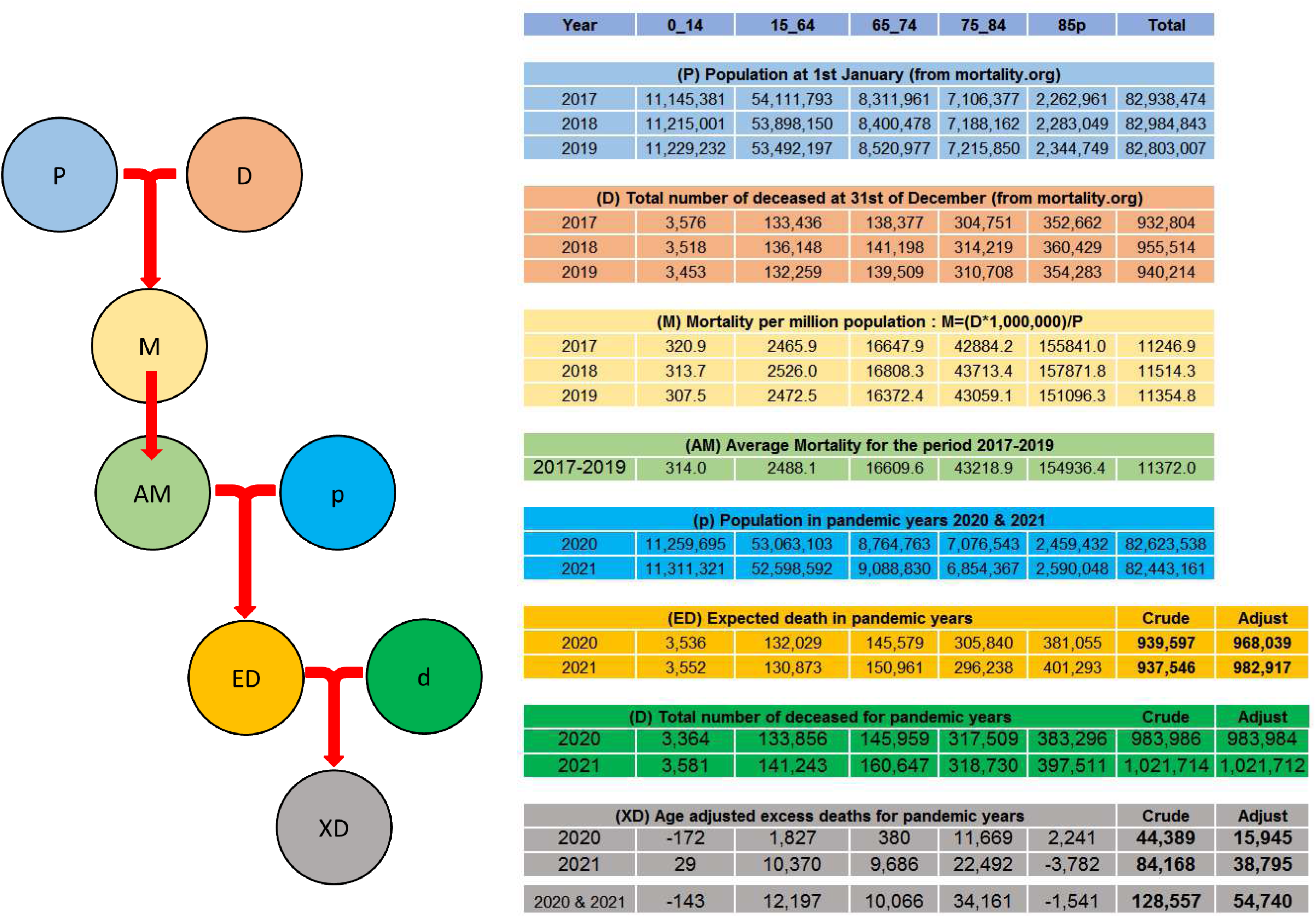
Schematic Representation of the Calculation of Age Adjusted Excess Death. Populations and total amounts of death on a single year are used to obtain the mortality values for the reference years (2017-2019) for each of the age strata. The average of this value is taken as the reference mortality for that strata. Expected deaths for a non COVID scenario for 2020 and 2021 are obtained from the population and reference mortality data for each strata. Excess deaths are calculated as the difference between the actual deaths and the expected deaths. Expected and excess deaths for the non age adjusted case are also reported for comparison. The table reports the real values for Germany as an example. This table is provided as Excel in the Supplement so that method can be easily used on other data.

Some clarifications are required for these analyses. First, on the calculation of the Weekly Population from the weekly Death Count and Weekly Mortality given in the MDH file stmf.csv. As Population = Death/Mortality, when Death = 0, Mortality = 0, so that Population is not defined. In these cases, we get the Population value from adjacent weeks with non-zero Death in the same year. Another problem is that some years, like 2015 & 2020 have an extra week 53, a leap week. This is because 52 weeks is 364 days whereas an average year is 365.25 days (a leap year every fourth year adds an extra day). Since mortality data are reported weekly, these years would get an apparent extra week of deaths. To correct this issue, we consider a standard year to be 365.25/7 = 52.1786 weeks and distribute weekly counts uniformly into years. Thus 2017 gets all its 52 week plus 0.1786 of week 1 of 2018. Year 2018 loses 0.1786 of week 1 and needs to get an additional 2*0.1786 of week 1 of 2019, and so on. In this way all the five years 2017 to 2021 are adjusted to have 52.1786 weeks.

We averaged the mortality values for each age-stratum over the reference years. We did not exclude any deaths or periods due to heat waves or other natural or man-made events, since it is subjective to arbitrate which ones should be excluded; moreover, we wanted to compare the COVID-19 pandemic with three recent years that had not raised any concerns about undue levels of deaths. We also repeated the excess death calculations using the same exact process but without considering age-strata.

We used population data, including also age-strata, from mortality.org.^34, 35^ for consistency across all evaluations in calculating excess deaths per million even though different evaluations had used different sources originally, e.g. WHO had used population data from the World Population Prospects 2019 with projections.^36^

### Countries considered in main comparison

We focused our primary comparison on countries that have excellent death registration, are high income, and include data with weekly deaths in the Human Mortality database. These countries have the most reliable evidence to allow proper excess death calculations and changes in deaths over time cannot be due to changes in death registration (e.g. either improvements in death registration during the pandemic as countries put more resources on capturing information or worsening of death registration during the pandemic due to the pandemonium and acute death peaks). We defined high-income countries by World Bank criteria.^37^ The Short Term Mortality Fluctuations file in the Human Mortality database includes detailed weekly deaths for the period 2017-2019 and also for the pandemic years 2020-2021 on 35 countries, all of which except Bulgaria and Russia are high-income, thus 33 countries were included in the main analyses. Of note, Canada and Australia did not have data for the entire 2 years of 2020-2021 and we performed calculations that cover the available time periods.

### Excess death metrics of interest

We calculated excess deaths for the full two years 2020-2021 for each country, E. We expressed them also as percentage above the expected deaths, E_p_ and as excess deaths per million E_m_. E.g. if 60,000 deaths happened in 2020-2021 in one country of 4,000,000 people and 50,000 were expected, the percentage E_P_ is (60,000-50,000)/50,000=12% and E_m_ is (60,000-50,000)/4,000,000=2,500 per million. As the main metric of interest, we used the ratio of estimated excess mortality E divided by the number of officially recorded COVID-19 deaths during 2020-2021, hence called the excess ratio R. E.g. R=1.20 means that the estimated excess mortality is 20% higher than the officially recorded COVID-19 deaths and R=0.90 means that the estimated excess mortality is 10% lower than the officially recorded COVID-19 deaths.

### Analyses

We calculated the total number of excess deaths across the eligible countries with each of the different calculation methods, and as compared with the total recorded COVID-19 deaths. We also calculated the Pearson correlation coefficients of excess death metrics across the eligible countries based on the different calculation methods (a full list of correlation coefficient estimates appears in Supplementary Table 5).

We noted in how many of the 33 eligible countries the different calculations of excess deaths had consistently R above 1 or R below 1 (i.e. they all agreed that excess deaths were more than the recorded COVID-19 deaths or they all agreed that excess deaths were less than the recorded COVID-19 deaths); and in how many countries had R values that were all within a range of 0.3 between the highest and lowest estimate (a higher range means that divergence in the excess death estimates exceeds 30% of the recorded COVID-19 deaths); and within a range of 1 (a higher range means that the divergence in the excess death estimates exceeds the number of recorded COVID-19 deaths itself).

For the remaining, non-eligible countries, we evaluated excess death metrics and their comparison across different evaluations as exploratory analyses.

All analyses were done independently by two analysts (ML and FZ) and then compared notes with arbitration (including discussion with the third author, JPAI) for any disagreements until both analysts obtained the same results.

### Data availability

All data are in the manuscript, tables, and supplementary tables and in the publicly available databases listed in Supplementary Links to Data

## Data Availability

All data are in the manuscript and in publicly available datasets

https://github.com/csblab

---

**Supplementary Table 1:** Excess deaths as percentage of the observed deaths in the 33 countries during 2020-2021.

| Country | HMD All-Cause Death in 2020 plus 2021 | Per eLife | Per Economist | Per Lancet | Per WHO | Per Levitt Age-Adjusted | Per Levitt Not Age-Adjusted |
| --- | --- | --- | --- | --- | --- | --- | --- |
| Australia | 277,603 | -4.2% | -3.4% | -6.5% | -5.1% | -5.2% | -0.8% |
| Austria | 180,363 | 8.5% | 9.4% | 10.1% | 6.6% | 7.2% | 8.5% |
| Belgium | 239,201 | 8.6% | 9.8% | 13.7% | 7.5% | 5.8% | 8.0% |
| Canada | 586,135 | 2.3% | 4.0% | 7.5% | 3.8% | 3.7% | 6.5% |
| Chile | 263,154 | 14.8% | 14.5% | 14.1% | 14.7% | 12.0% | 17.1% |
| Croatia | 119,871 | 14.0% | 16.0% | 19.1% | 14.3% | 10.2% | 13.4% |
| Czechia | 269,137 | 15.4% | 16.3% | 18.2% | 13.8% | 12.7% | 16.1% |
| Denmark | 111,772 | 0.8% | 2.2% | 9.3% | 3.3% | -2.8% | 2.1% |
| Estonia | 34,559 | 9.2% | 10.9% | 16.3% | 9.8% | 7.7% | 9.7% |
| Finland | 112,800 | 2.4% | 4.0% | 7.8% | 2.5% | -0.6% | 3.9% |
| France | 1,297,407 | 6.1% | 7.5% | 11.9% | 6.3% | 4.5% | 7.5% |
| Germany | 2,005,701 | 4.4% | 5.6% | 10.1% | 9.7% | 2.7% | 6.4% |
| Greece | 274,725 | 8.8% | 9.2% | 9.2% | 7.1% | 7.5% | 10.8% |
| Hungary | 296,496 | 12.1% | 14.1% | 18.1% | 12.3% | 9.4% | 12.2% |
| Iceland | 4,640 | 1.1% | -0.8% | -6.8% | -0.2% | -3.0% | 0.2% |
| Israel | 99,437 | 7.2% | 8.0% | 9.3% | 6.2% | 3.2% | 5.5% |
| Italy | 1,454,193 | 11.5% | 13.1% | 17.8% | 11.1% | 8.0% | 11.4% |
| Latvia | 63,088 | 11.1% | 12.4% | 19.7% | 12.2% | 9.6% | 11.1% |
| Lithuania | 90,523 | 17.7% | 19.2% | 22.1% | 19.1% | 12.5% | 13.6% |
| Luxembourg | 9,106 | 0.6% | 3.4% | 11.8% | 0.8% | 1.2% | 1.9% |
| Netherlands | 339,242 | 8.4% | 9.7% | 13.4% | 8.6% | 5.3% | 9.4% |
| New Zealand | 67,586 | -4.1% | -3.8% | -1.3% | -4.0% | -6.1% | -2.7% |
| Norway | 82,491 | 1.3% | 2.4% | 0.9% | -0.1% | -3.6% | -0.2% |
| Poland | 998,284 | 15.8% | 17.2% | 21.4% | 15.8% | 15.0% | 18.3% |
| Portugal | 248,658 | 8.3% | 9.9% | 16.2% | 8.2% | 6.5% | 10.3% |
| Slovakia | 131,782 | 18.3% | 19.4% | 19.3% | 18.5% | 14.2% | 18.0% |
| Slovenia | 47,090 | 10.5% | 11.7% | 14.8% | 11.9% | 8.4% | 11.9% |
| South Korea | 621,862 | 1.2% | 1.1% | 0.7% | 1.0% | -4.9% | 5.4% |
| Spain | 948,016 | 10.9% | 12.2% | 17.1% | 11.0% | 7.2% | 10.1% |
| Sweden | 184,326 | 5.4% | 6.5% | 9.8% | 6.1% | -0.2% | 2.0% |
| Switzerland | 146,969 | 7.8% | 9.2% | 10.5% | 5.6% | 3.8% | 6.9% |
| United Kingdom | 1,353,941 | 10.1% | 11.0% | 12.5% | 11.0% | 6.4% | 9.3% |
| United States | 6,849,500 | 14.0% | 14.9% | 16.5% | 13.6% | 12.7% | 16.3% |
| TOTAL OR AVERAGES | 19,809,658 | 7.9% | 9.0% | 11.7% | 8.0% | 5.2% | 8.5% |

**Supplementary Table 2:** Excess deaths per million population in the 33 countries during 2020-2021.

| Country | Mean 2020<br>& 2021<br>Population<br>(millions)<br>from<br>HMD** | Excess Death<br>per eLife | Excess Death<br>per Economist | Excess Death<br>per Lancet | WHO | Excess Death<br>per Levitt<br>Age-Adjusted | Excess Death<br>per Levitt Not<br>Age-Adjusted |
| --- | --- | --- | --- | --- | --- | --- | --- |
| Australia | 24.547 | -474 | -387 | -737 | -581 | -589 | -86 |
| Austria | 8.935 | 1,708 | 1,889 | 2,048 | 1,336 | 1,456 | 1,717 |
| Belgium | 11.494 | 1,793 | 2,033 | 2,854 | 1,559 | 1,214 | 1,656 |
| Canada | 36.108 | 373 | 652 | 1,210 | 610 | 605 | 1,051 |
| Chile | 17.960 | 2,166 | 2,121 | 2,071 | 2,155 | 1,762 | 2,507 |
| Croatia | 4.051 | 4,154 | 4,736 | 5,653 | 4,240 | 3,013 | 3,962 |
| Czechia | 10.730 | 3,866 | 4,095 | 4,576 | 3,452 | 3,176 | 4,032 |
| Denmark | 5.864 | 156 | 418 | 1,774 | 634 | -538 | 408 |
| Estonia | 1.332 | 2,381 | 2,833 | 4,227 | 2,533 | 2,008 | 2,512 |
| Finland | 5.548 | 480 | 806 | 1,583 | 515 | -129 | 783 |
| France | 65.467 | 1,205 | 1,488 | 2,368 | 1,250 | 882 | 1,479 |
| Germany | 82.533 | 1,072 | 1,372 | 2,460 | 2,363 | 663 | 1,558 |
| Greece | 10.711 | 2,257 | 2,359 | 2,371 | 1,811 | 1,915 | 2,759 |
| Hungary | 9.762 | 3,668 | 4,273 | 5,511 | 3,739 | 2,849 | 3,697 |
| Iceland | 0.362 | 138 | -97 | -867 | -28 | -391 | 30 |
| Israel | 9.293 | 775 | 857 | 999 | 665 | 344 | 583 |
| Italy | 59.630 | 2,814 | 3,201 | 4,343 | 2,697 | 1,940 | 2,790 |
| Latvia | 1.906 | 3,662 | 4,119 | 6,506 | 4,023 | 3,172 | 3,685 |
| Lithuania | 2.802 | 5,713 | 6,208 | 7,138 | 6,157 | 4,027 | 4,380 |
| Luxembourg | 0.635 | 90 | 494 | 1,685 | 109 | 171 | 270 |
| Netherlands | 17.479 | 1,630 | 1,889 | 2,603 | 1,671 | 1,028 | 1,832 |
| New Zealand | 5.013 | -556 | -512 | -174 | -534 | -821 | -364 |
| Norway | 5.408 | 204 | 367 | 137 | -18 | -554 | -34 |
| Poland | 38.482 | 4,086 | 4,465 | 5,561 | 4,094 | 3,891 | 4,741 |
| Portugal | 10.323 | 2,003 | 2,376 | 3,914 | 1,981 | 1,578 | 2,480 |
| Slovakia | 5.480 | 4,403 | 4,660 | 4,635 | 4,438 | 3,406 | 4,341 |
| Slovenia | 2.103 | 2,355 | 2,612 | 3,319 | 2,655 | 1,876 | 2,671 |
| South Korea | 51.631 | 146 | 135 | 90 | 122 | -587 | 647 |
| Spain | 47.511 | 2,168 | 2,435 | 3,410 | 2,188 | 1,446 | 2,020 |
| Sweden | 10.408 | 954 | 1,151 | 1,739 | 1,081 | -35 | 352 |
| Switzerland | 8.688 | 1,311 | 1,558 | 1,784 | 949 | 649 | 1,167 |
| United Kingdom | 67.145 | 2,037 | 2,217 | 2,517 | 2,218 | 1,300 | 1,872 |
| United States | 329.995 | 2,912 | 3,084 | 3,424 | 2,826 | 2,640 | 3,382 |
| TOTAL OR<br>AVERAGE | 969.336 | 1,868 | 2,118 | 2,749 | 1,906 | 1,314 | 1,966 |

**Supplementary Table 3:** Excess deaths in 42 countries with available calculations from eLife, Economist, Lancet, and WHO.

| Country | 2021<br>Population<br>(millions)<br>from WPR | Reported<br>COVID-19<br>Deaths<br>(OWID) | Excess<br>Death per<br>eLife | Excess<br>Death per<br>Economist | Excess<br>Death per<br>Lancet | Excess<br>Death per<br>WHO |
| --- | --- | --- | --- | --- | --- | --- |
| Albania | 2.873 | 3,217 | 14,655 | 15,514 | 17,300 | 12,688 |
| Armenia | 2.968 | 7,972 | 19,882 | 14,023 | 20,600 | 19,663 |
| Azerbaijan | 10.223 | 8,358 | 39,829 | 39,505 | 53,500 | 57,006 |
| Bolivia | 11.833 | 19,680 | 55,330 | 54,059 | 161,000 | 88,029 |
| Bosnia and Herzegovina | 3.263 | 13,442 | 19,123 | 21,019 | 20,900 | 15,719 |
| Brazil | 213.993 | 619,334 | 698,346 | 696,006 | 792,000 | 681,266 |
| Bulgaria | 6.897 | 30,955 | 58,061 | 61,754 | 82,500 | 57,495 |
| Colombia | 51.266 | 129,942 | 165,781 | 165,571 | 179,000 | 164,745 |
| Cyprus | 1.216 | 638 | 783 | 725 | 809 | 1,011 |
| Ecuador | 17.888 | 33,681 | 66,620 | 67,917 | 112,000 | 80,867 |
| Egypt | 104.258 | 21,752 | 275,979 | 296,179 | 265,000 | 251,101 |
| Guatemala | 18.250 | 16,107 | 43,111 | 43,389 | 51,100 | 49,393 |
| Iran | 85.029 | 131,606 | 251,283 | 258,514 | 274,000 | 232,153 |
| Ireland | 4.983 | 5,912 | 2,954 | 3,900 | 1,170 | 2,920 |
| Japan | 126.051 | 18,389 | -13,147 | 15,873 | 111,000 | -19,469 |
| Kazakhstan | 18.995 | 18,211 | 80,686 | 81,946 | 49,000 | 76,214 |
| Kyrgyzstan | 6.628 | 2,802 | 14,532 | 13,906 | 22,600 | 12,313 |
| Lebanon | 6.769 | 9,119 | 15,149 | 14,111 | 36,100 | 18,518 |
| Malta | 0.443 | 476 | 405 | 500 | 735 | 477 |
| Mauritius | 1.273 | 786 | 1,097 | 1,315 | 659 | 942 |
| Mexico | 130.262 | 299,428 | 606,476 | 616,288 | 798,000 | 626,219 |
| Moldova | 4.024 | 10,275 | 15,702 | 16,491 | 16,500 | 18,158 |
| Monaco | 0.040 | 38 | 134 | 164 | 53 | 64 |
| Mongolia | 3.329 | 2,059 | 721 | 1,088 | 5,600 | 6 |
| Montenegro | 0.628 | 2,411 | 2,632 | 3,534 | 4,080 | 3,910 |
| North Macedonia | 2.083 | 7,960 | 14,242 | 15,407 | 20,200 | 15,358 |
| Oman | 5.223 | 4,116 | 4,205 | 4,332 | 12,300 | 11,502 |
| Panama | 4.382 | 7,428 | 7,498 | 7,375 | 10,200 | 7,643 |
| Paraguay | 7.220 | 16,624 | 19,863 | 19,481 | 22,500 | 19,817 |
| Peru | 33.359 | 202,690 | 217,194 | 209,350 | 349,000 | 289,667 |
| Philippines | 111.047 | 51,504 | 204,234 | 223,349 | 184,000 | 185,255 |
| Qatar | 2.931 | 618 | 920 | 913 | 1,560 | 1,538 |
| Romania | 19.128 | 58,752 | 108,159 | 115,973 | 119,000 | 106,912 |
| Russia | 145.912 | 302,671 | 1,080,748 | 1,090,178 | 1,070,000 | 1,072,326 |
| San Marino | 0.034 | 100 | 133 | 135 | 118 | 169 |
| Serbia | 8.698 | 12,714 | 50,973 | 52,167 | 37,400 | 55,643 |
| Singapore | 5.897 | 828 | 1,766 | 1,156 | -1,770 | 1,477 |
| South Africa | 60.042 | 91,145 | 246,097 | 243,575 | 302,000 | 238,670 |
| Thailand | 69.951 | 21,698 | 63,120 | 60,679 | 35,200 | 15,303 |
| Ukraine | 43.467 | 102,088 | 178,896 | 191,797 | 181,000 | 198,226 |
| Uruguay | 3.485 | 6,170 | 3,023 | 3,213 | 9,820 | 3,108 |
| Uzbekistan | 33.936 | 1,485 | 35,729 | 34,662 | 69,100 | 44,888 |
| TOTALS | 1390.176 | 2,295,181 | 4,672,924 | 4,777,033 | 5,496,834 | 4,718,910 |

**Supplementary Table 4:**
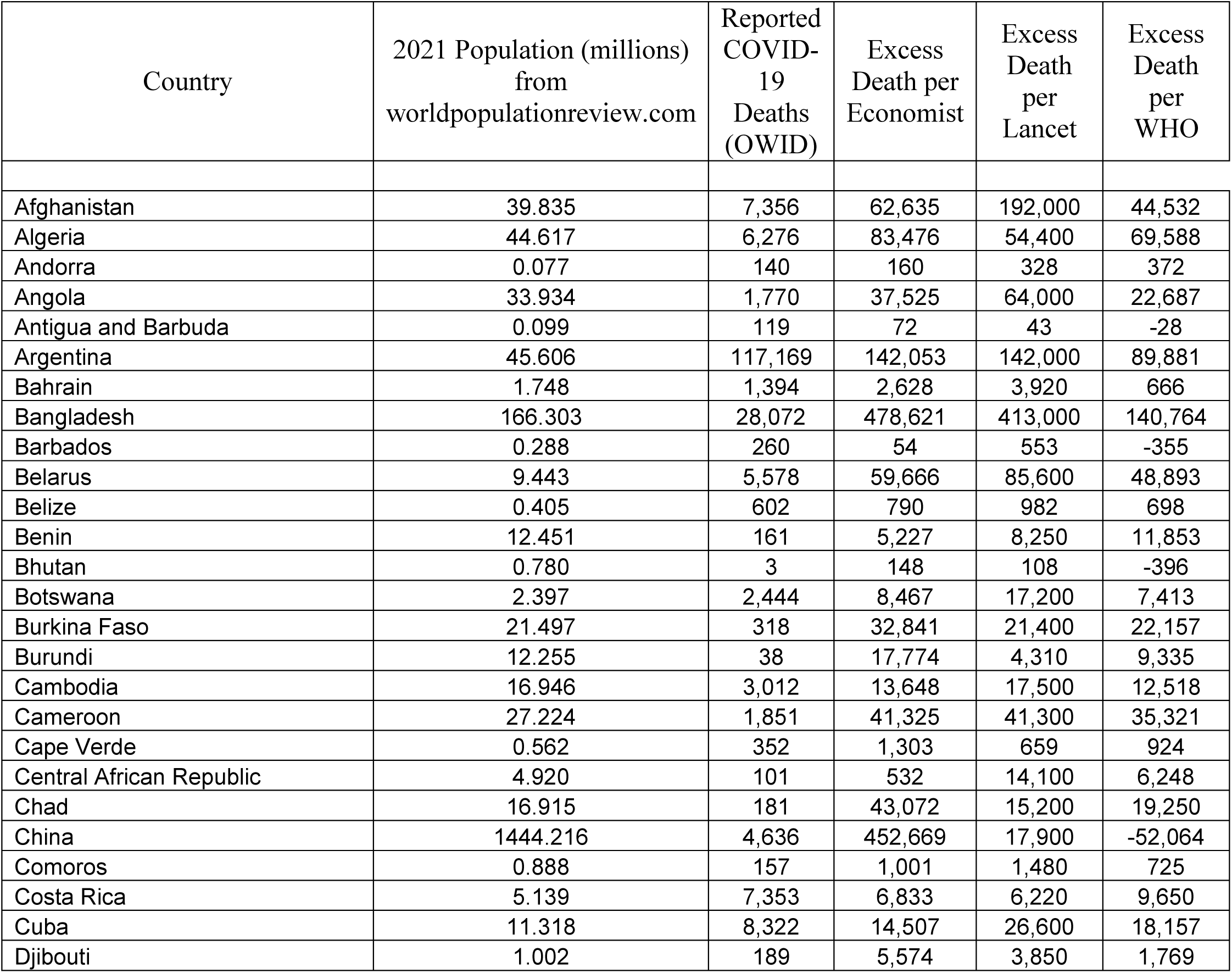
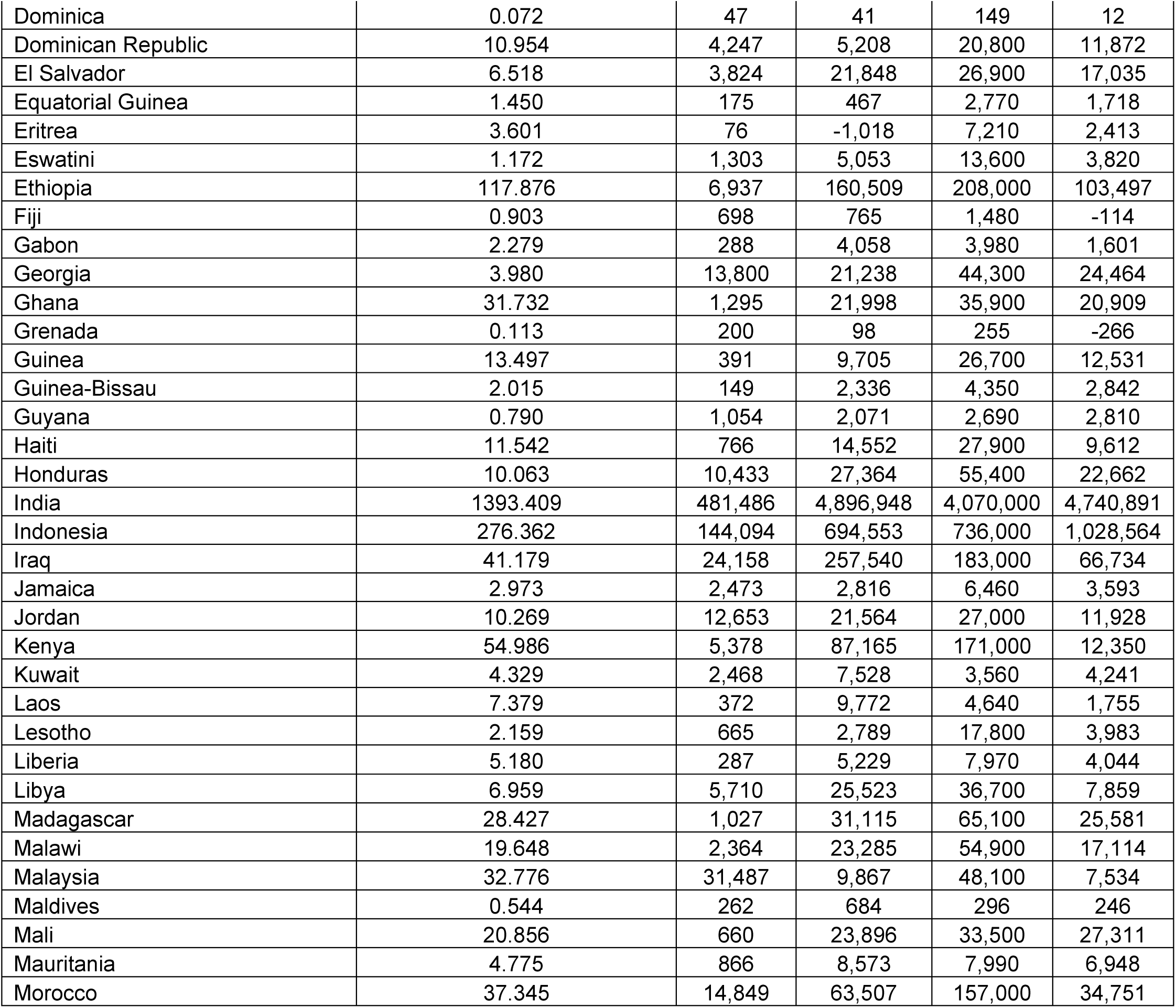
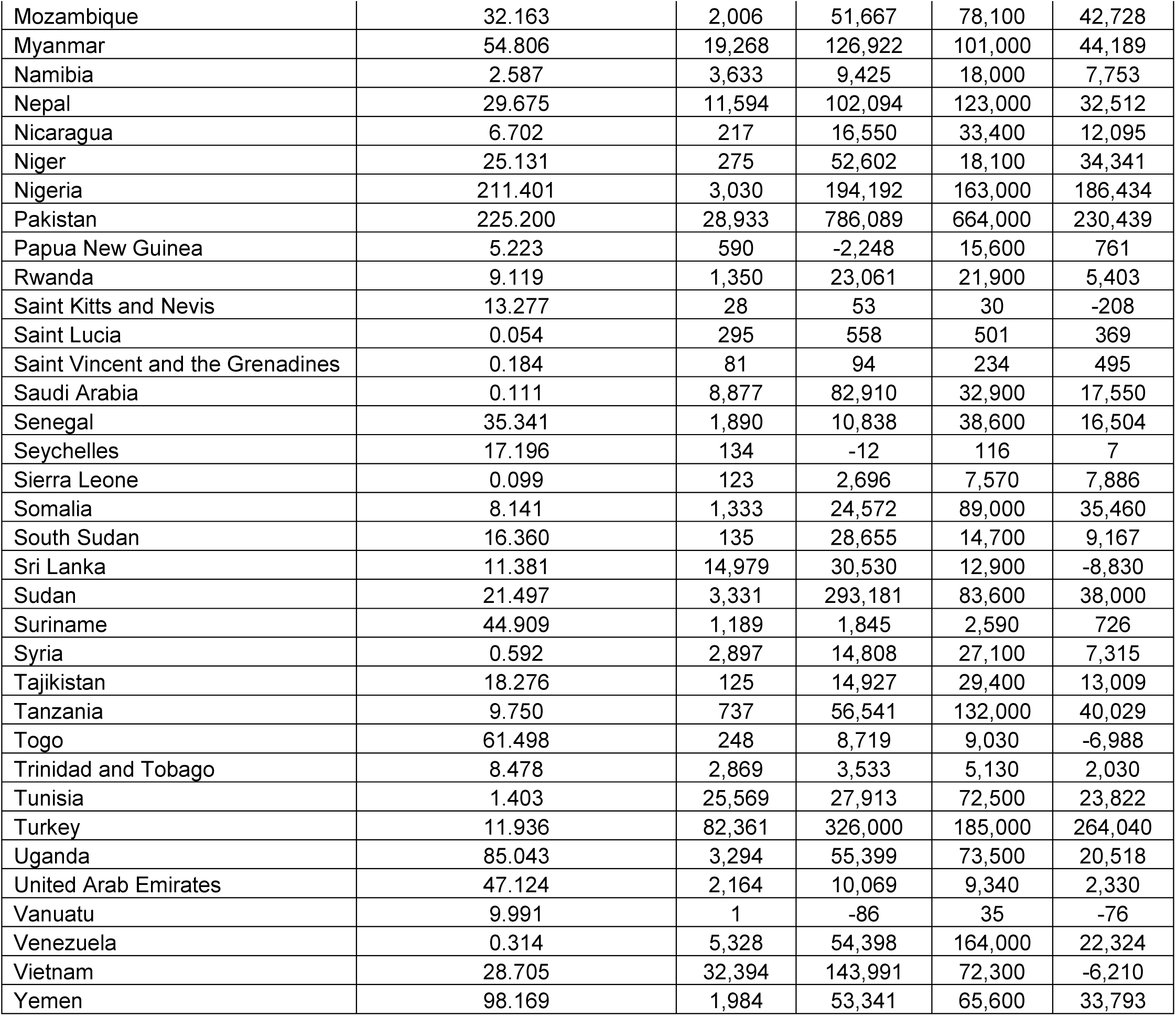
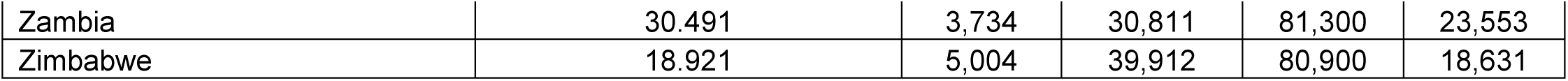
Excess deaths in 98 countries with available calculations from Economist, Lancet and WHO.

**Supplementary Table 5:**
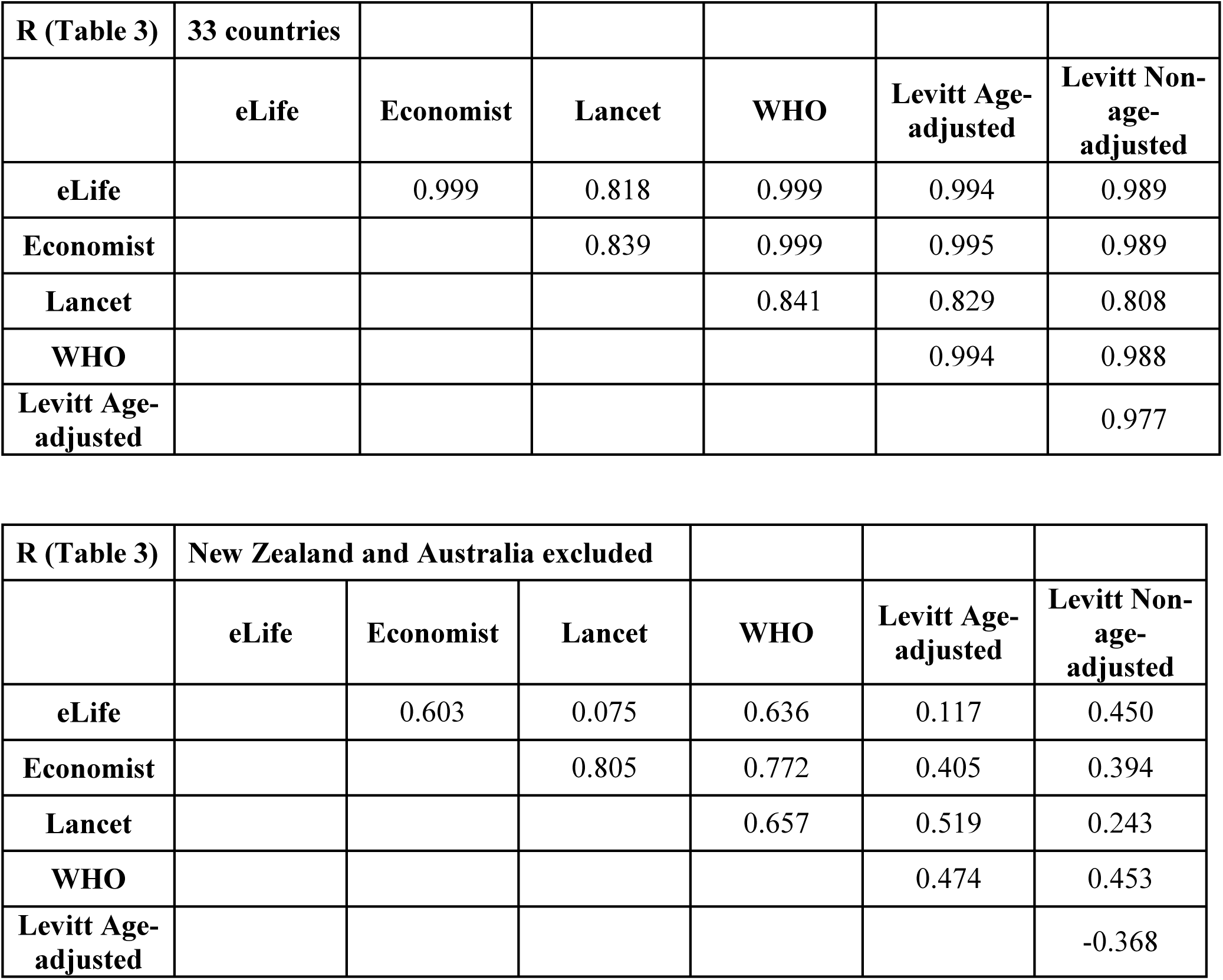
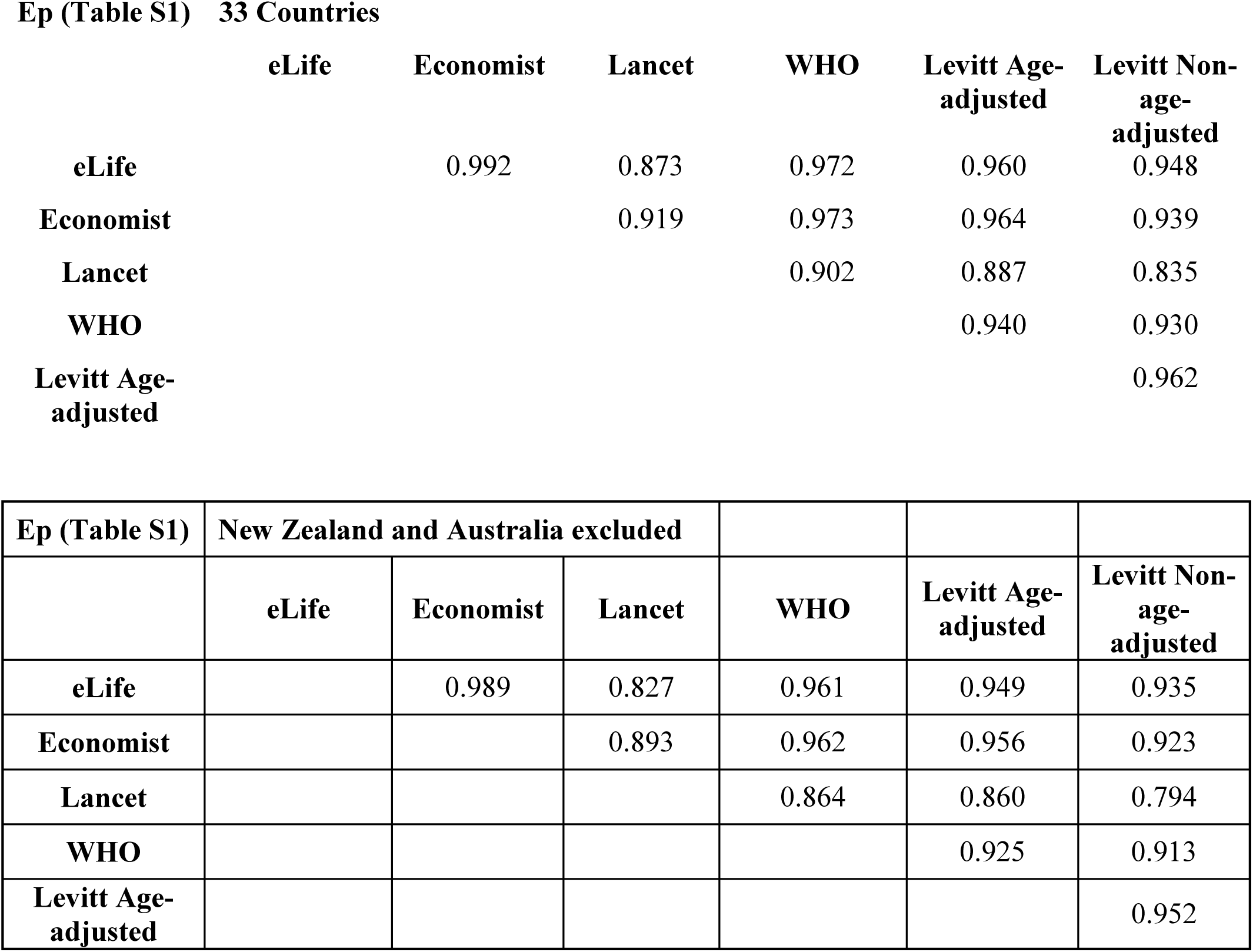
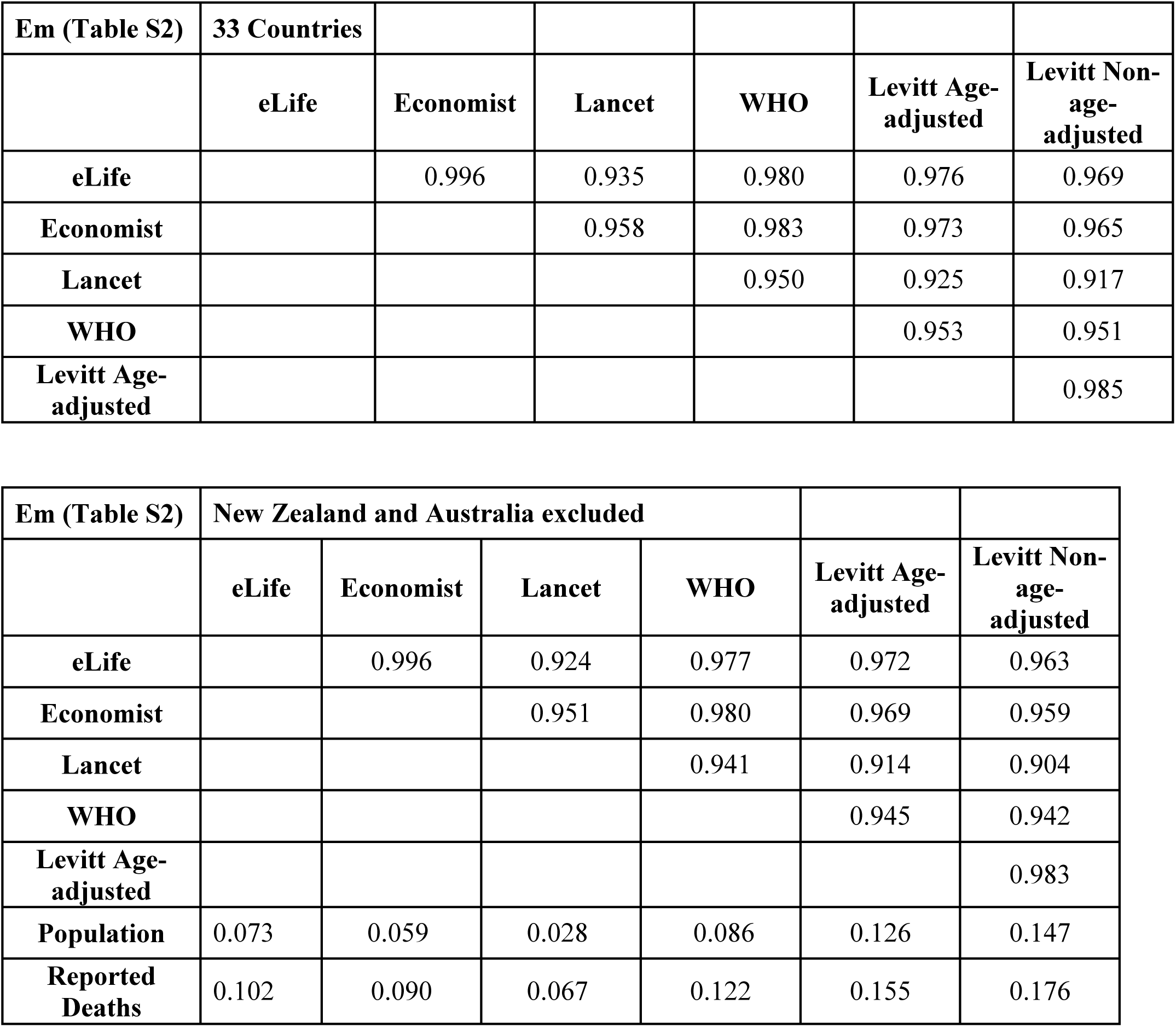
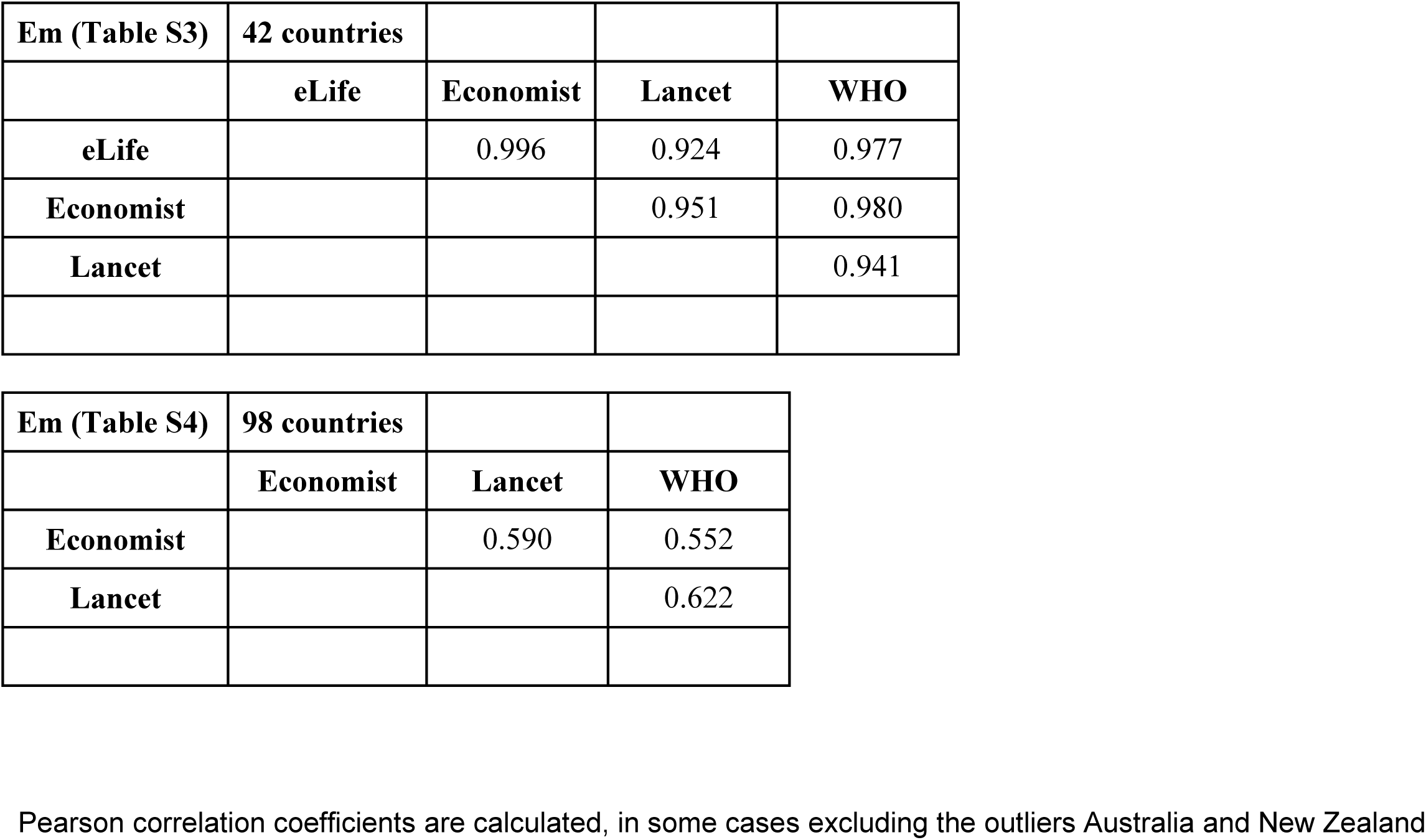
Correlation coefficients for various excess death metrics (as shown in Table 3, supplementary Table 1, supplementary Table 2, supplementary Table 3, and supplementary Table 4) between the different evaluations.

| **R (Table 3)** | **33 countries** |  |  |  |  |  |
| --- | --- | --- | --- | --- | --- | --- |
|  | **eLife** | **Economist** | **Lancet** | **WHO** | **Levitt Age-adjusted** | **Levitt Non-age-adjusted** |
| **eLife** |  | 0.999 | 0.818 | 0.999 | 0.994 | 0.989 |
| **Economist** |  |  | 0.839 | 0.999 | 0.995 | 0.989 |
| **Lancet** |  |  |  | 0.841 | 0.829 | 0.808 |
| **WHO** |  |  |  |  | 0.994 | 0.988 |
| **Levitt Age-adjusted** |  |  |  |  |  | 0.977 |

| **R (Table 3)** | **New Zealand and Australia excluded** |  |  |  |  |  |
| --- | --- | --- | --- | --- | --- | --- |
|  | **eLife** | **Economist** | **Lancet** | **WHO** | **Levitt Age-adjusted** | **Levitt Non-age-adjusted** |
| **eLife** |  | 0.603 | 0.075 | 0.636 | 0.117 | 0.450 |
| **Economist** |  |  | 0.805 | 0.772 | 0.405 | 0.394 |
| **Lancet** |  |  |  | 0.657 | 0.519 | 0.243 |
| **WHO** |  |  |  |  | 0.474 | 0.453 |
| **Levitt Age-adjusted** |  |  |  |  |  | -0.368 |

**Ep (Table S1) 33 Countries**
|  | **eLife** | **Economist** | **Lancet** | **WHO** | **Levitt Age-**<br>**adjusted** | **Levitt Non-**<br>**age-**<br>**adjusted** |
| --- | --- | --- | --- | --- | --- | --- |
| **eLife** |  | 0.992 | 0.873 | 0.972 | 0.960 | 0.948 |
| **Economist** |  |  | 0.919 | 0.973 | 0.964 | 0.939 |
| **Lancet** |  |  |  | 0.902 | 0.887 | 0.835 |
| **WHO** |  |  |  |  | 0.940 | 0.930 |
| **Levitt Age-**<br>**adjusted** |  |  |  |  |  | 0.962 |

| **Ep (Table S1)** | **New Zealand and Australia excluded** |  |  |  |  |  |
| --- | --- | --- | --- | --- | --- | --- |
|  | **eLife** | **Economist** | **Lancet** | **WHO** | **Levitt Age-**<br>**adjusted** | **Levitt Non-**<br>**age-**<br>**adjusted** |
| **eLife** |  | 0.989 | 0.827 | 0.961 | 0.949 | 0.935 |
| **Economist** |  |  | 0.893 | 0.962 | 0.956 | 0.923 |
| **Lancet** |  |  |  | 0.864 | 0.860 | 0.794 |
| **WHO** |  |  |  |  | 0.925 | 0.913 |
| **Levitt Age-**<br>**adjusted** |  |  |  |  |  | 0.952 |

| **Em (Table S2)** | **33 Countries** |  |  |  |  |  |
| --- | --- | --- | --- | --- | --- | --- |
|  | **eLife** | **Economist** | **Lancet** | **WHO** | **Levitt Age-adjusted** | **Levitt Non-age-adjusted** |
| **eLife** |  | 0.996 | 0.935 | 0.980 | 0.976 | 0.969 |
| **Economist** |  |  | 0.958 | 0.983 | 0.973 | 0.965 |
| **Lancet** |  |  |  | 0.950 | 0.925 | 0.917 |
| **WHO** |  |  |  |  | 0.953 | 0.951 |
| **Levitt Age-adjusted** |  |  |  |  |  | 0.985 |

| **Em (Table S2)** | **New Zealand and Australia excluded** |  |  |  |  |  |
| --- | --- | --- | --- | --- | --- | --- |
|  | **eLife** | **Economist** | **Lancet** | **WHO** | **Levitt Age-adjusted** | **Levitt Non-age-adjusted** |
| **eLife** |  | 0.996 | 0.924 | 0.977 | 0.972 | 0.963 |
| **Economist** |  |  | 0.951 | 0.980 | 0.969 | 0.959 |
| **Lancet** |  |  |  | 0.941 | 0.914 | 0.904 |
| **WHO** |  |  |  |  | 0.945 | 0.942 |
| **Levitt Age-adjusted** |  |  |  |  |  | 0.983 |
| **Population** | 0.073 | 0.059 | 0.028 | 0.086 | 0.126 | 0.147 |
| **Reported Deaths** | 0.102 | 0.090 | 0.067 | 0.122 | 0.155 | 0.176 |

|  |  |  |  |  |
| --- | --- | --- | --- | --- |
| **Em (Table S3)** | **42 countries** |  |  |  |
|  | **eLife** | **Economist** | **Lancet** | **WHO** |
| **eLife** |  | 0.996 | 0.924 | 0.977 |
| **Economist** |  |  | 0.951 | 0.980 |
| **Lancet** |  |  |  | 0.941 |

|  |  |  |  |
| --- | --- | --- | --- |
| **Em (Table S4)** | **98 countries** |  |  |
|  | **Economist** | **Lancet** | **WHO** |
| **Economist** |  | 0.590 | 0.552 |
| **Lancet** |  |  | 0.622 |

Pearson correlation coefficients are calculated, in some cases excluding the outliers Australia and New Zealand

## Supplementary Links to Data

HMD

1-May-2022

https://www.mortality.org/Public/STMF/Outputs/stmf.csv

Use this for 33 set Populations, Total deaths & Excess deaths in age-bands

OWID for COVID-19 reported cases and excess death by eLife method. 22-Apr-2022

https://covid.ourworldindata.org/data/owid-covid-data.csv

Use this for eLife & Reported COVID-19 Cumulative count of Death to date of Completion

WPR for estimate Deaths 14-Apr-2022

https://worldpopulationreview.com/country-rankings/death-rate-by-country Use rateGHDE?

Expected death is 2 years (rateGHDE X pop2021) csvData.csv

https://worldpopulationreview.com/countries Use pop2021 in column [D]

csvData.csv

World Bank 14-Apr-2022

https://data.worldbank.org/indicator/SP.DYN.CDRT.IN?end=2017&start=2017&view=bar

**API_SP.DYN.CDRT.IN_DS2_en_csv_v2_3932073.csv**

Lancet

5-Apr-2022

https://ghdx.healthdata.org/sites/default/files/record-attached-files/IHME_EM_COVID_19_2020_2021_DATA_Y2022M03D10.CSV

Use for Lancet Reported deaths and Excess Death for 2020 &; 2021

Economist 3-Apr-2022

https://ourworldindata.org/excess-mortality-covid#estimated-excess-mortality-from-the-economist

**excess-deaths-cumulative-economist-single-entity.csv**

## Notes

### Competing Interest Statement

The authors have declared no competing interest.

### Funding Statement

NIH R35 GM122543

